# Artificial Intelligence algorithm for real-time detection and counting of *Trypanosoma cruzi* parasites using smartphone microscopy

**DOI:** 10.1101/2025.03.03.25323227

**Authors:** Lin Lin, Ana Valeria Solano, Fabiola Gonzales, Mary Cruz Torrico, Daniel Illanes, Nuria Díez, David Bermejo-Peláez, Elena Dacal, Ramón Vallés-López, Lucia Pastor, Roberto Mancebo-Martín, María Jesús Ledesma-Carbayo, Miguel Luengo-Oroz, Jose M. Rubio, Maria Flores-Chavez

## Abstract

Chagas disease affects 6–7 million people worldwide and causes approximately 12,000 deaths annually. Diagnostic methods vary by disease stage, with serological tests commonly used in the chronic phase, while microscopy and molecular techniques like PCR and LAMP are employed in the acute phase. While microscopy remains the most accessible tool in resource constrained settings, its effectiveness depends on skilled personnel, creating diagnostic bottlenecks.

To overcome these limitations, we developed a portable, smartphone-integrated AI system for real-time *Trypanosoma cruzi* detection in microscopy images. The platform combines a 3D-printed microscope adapter which aligns the smartphone camera with microscope ocular to digitize images, telemedicine-enabled annotation workflows, and lightweight AI models (SSD-MobileNetV2, YOLOv8) deployed on smartphone for real-time analysis. Trained on a diverse dataset of human samples (478 images from 20 samples), including thick/thin blood smears and cerebrospinal fluid) and murine thin smears (570 images from 33 samples), the SSD-MobileNetV2 model achieved 86% precision, 87% recall, and 86.5% F1-score on human samples, demonstrating robust performance across variable imaging conditions.

This system enables rapid, accurate parasite detection in field settings without advanced infrastructure, addressing critical gaps in early diagnosis and monitoring. Its modular design allows adaptation to other pathogens and cellular structures, offering a scalable solution for neglected tropical disease diagnostics. By bridging AI innovation with microscopy, this approach holds promise for advancing equitable healthcare delivery in endemic regions and aligning with global health priorities.

**Author Summary:** Chagas disease is a life-threatening illness affecting millions, primarily in Latin America, where access to advanced laboratory equipment and trained specialists is limited. One method of diagnosis is microscopic examination of blood or cerebrospinal fluid samples, it provides immediate results without requiring complex facilities, but its effectiveness depends on the expertise of trained microscopists.

We developed a simple, low-cost tool that combines a smartphone, a light microscope, and artificial intelligence (AI) to assist the diagnosis. By attaching a smartphone to a microscope with a 3D-printed adapter, users analyze microscopy images with real-time AI assistance, detecting the parasites causing Chagas disease.

Our results show that this approach is accurate and could be useful for regions with limited healthcare infrastructure. It reduces reliance on specialized training and expensive equipment, helping communities diagnose Chagas faster.

Beyond Chagas, this approach can be adapted to detect other diseases, offering a versatile tool for fighting neglected tropical illnesses. By bridging gaps in healthcare access, we hope to empower frontline workers and contribute to global efforts to reduce the burden of these diseases on vulnerable populations.

## Introduction

Chagas disease, also known as American trypanosomiasis, is a life-threatening illness caused by infection with the protozoan parasite *Trypanosoma cruzi*. It affects about 6-7 million people worldwide and it’s endemic in 21 Latin American countries, leading to approximately 12,000 deaths every year [1]. The *T. cruzi* is mainly transmitted by contact with feces/urine of triatomine bugs, but can also be transmitted via consumption of contaminated food or beverages, congenital, transfusional, organ transplantation and laboratory accidents.

The disease presents two phases, during the initial acute phase, a high number of parasites circulates in the blood, but the symptoms are mild or absent, including fever, headache, enlarged lymph glands, etc. In particular situations, myocarditis and meningoencephalitis can be observed. The acute phase is followed by the chronicle phase, with the parasites hidden in the hearts and digestive muscle, causing cardiac, digestive, or neurological disorder, leading to death [2]. In immunosuppressive conditions such as HIV co-infection, parasitemia increases to levels similar to those in the acute phase, and may cross the blood-brain barrier, causing cerebral chagoma [3,4].

The diagnosis of Chagas disease can vary depending on the stage of the disease (acute or chronic) and the resources available. During the chronicle phase, the diagnosis is mainly performed using serological tests, including Enzyme-Linked Immunosorbent Assay (ELISA) and Indirect Immunofluorescence Assay (IFA) due to their ability to detect antibodies against *T. cruzi* [5,6]. In contrast, the acute phase is characterized by high parasitemia, enabling direct parasite detection through microscopy or molecular techniques, including PCR and loop-mediated isothermal amplification (LAMP) [7]. While molecular methods offer rapid and highly sensitive detection of the parasitaemia, their reliance on costly equipment, specialized training, and advanced laboratory infrastructure limits their utility in resource-constrained settings. Microscopy, on the other hand, remains a cornerstone of acute-phase diagnosis due to its ability to visualize *T. cruzi* trypomastigotes in fresh blood or cerebrospinal fluid smears (CSF) samples or stained smears. This method provides immediate results without requiring complex facilities, though its effectiveness depends on the expertise of trained microscopists to ensure high clinical sensitivity and specificity [7–9].

Microscopy is particularly valuable in regions where Chagas disease overlaps with other endemic infections, such as malaria. For example, in the Brazilian western Amazon—a region with high malaria prevalence and sporadic Chagas outbreaks linked to oral transmission via contaminated acai juice—febrile illnesses are often presumptively attributed to *Plasmodium* infections [10]. However, microscopy enables the incidental identification of *T. cruzi* during routine blood smear analysis for malaria, as the trypomastigotes exhibit distinct morphological features distinguishable from malaria parasites [11]. This dual diagnostic capacity has promoted initiatives in the Brazilian Amazon to train microscopy personnel in recognizing both pathogens, enhancing early detection and treatment in areas with limited access to advanced diagnostic tools [11].

Recent advancements in artificial intelligence (AI) are poised to address key limitations of traditional microscopy while amplifying its strengths. AI-driven tools are increasingly being applied to medical image analysis [12,13] In addition to radiology, AI also holds great potential for microscopy image analysis, including pathology [14,15], hematology [16,17], and parasitology [18,19], such as malaria [20–22], Schistosoma [23–25], soil-transmitted helminthiasis [26–29] or filariasis [30]. For Chagas disease, AI models have shown promise in detecting *T. cruzi* trypomastigotes in blood smears with high accuracy. Researchers have employed various image processing techniques combined with AI to detect and quantify *T. cruzi* parasites. The algorithms developed for this purpose can be classified into three main categories: the first category involves traditional image processing methods to extract relevant features, which are then analyzed using machine learning algorithms; the second category relies on convolutional neural networks (CNNs) for automatic feature extraction from raw images, enabling the classification of images as infected or non-infected; and the third category utilizes CNN-based segmentation methods that not only determine infection status but also provide detailed information on the distribution of parasites within a sample [31–33].

In the first category, Soberanis et al. proposed an approach where the image is first segmented and then classified using the k-nearest neighbor algorithm [34]. Uc-Cetina et al. developed a parasite detection method using AdaBoost and Support Vector Machine (SVM), which achieved a sensitivity of 100% and a specificity of 93.25% [35]. Both studies used the same dataset, which consisted of blood samples from infected mice, digitized at 100x magnification, and cropped to 256×256 pixels. Another notable work within this category is by Morais et al., who introduced a system that combines graph-based segmentation with random forest classification. In this method, images acquired via mobile phones were segmented using a graph-based technique, with areas of interest cropped into 100×100 patches, which were then classified using a random forest algorithm. This system achieved a precision (positive predictive value) of 88.6% and a recall (sensitivity) of 90.5% [31].

The second category includes the work of Pereira et al., who proposed the first image classification algorithm using CNNs to identify whether a 224×224 pixel patch contains *T. cruzi* parasites. Their model, based on MobileNet and trained on 331 image patches with testing on 214 patches, achieved a precision of 91.8% and a recall of 50.5% [32]. Similarly, Jung et al. utilized CNN ResNet18 for classifying 224×224 pixel patch images, achieving a sensitivity of 70.6% and specificity of 65.1% [36].

In the third category, segmentation using CNNs has been a focal point. Ojeda et al. proposed a U-Net based approach for segmenting images of 512×512 pixels, which resulted in a precision of 63.04% and a recall of 87.02% [37]. In another study, Sanchez-Patiño et al. applied U-Net for the segmentation of *T. cruzi* parasites in histopathological images, achieving a binary accuracy of 98.19% [33].

Despite the promising advancements in applying AI and image processing techniques for diagnosing Chagas disease, several drawbacks and limitations persist in these works that must be addressed to improve their clinical utility and reliability. One significant limitation of those algorithms is the ability to execute the algorithm at the point of care in almost real time. For algorithms of the first and second category, they use small patches covering a small area of the sample preparation, for a single field of view, several patches are generated, leading to computational inefficiency. On the other hand, the segmentation algorithms usually have high computational complexity, posing a challenge on the deployment of the algorithm on resource-constrained settings.

Building upon the challenges of existing methods, in this work we propose an end to end pipeline to create and deploy an AI algorithm for detection of the *T. cruzi* parasites. The pipeline includes image digitization with smartphones coupled to a light microscope through a 3D-printed device, image labeling on the telemedicine platform, AI algorithm training and the deployment of the AI algorithm on the smartphone to assist the *T. cruzi* parasite count in real time. The proposed algorithm is lightweight and can be deployed in the field, requiring few computing resources, making it an ideal tool for early detection and monitoring of Chagas disease in endemic regions, contributing to the achievement of the World Health Organization targets of eradication and control of Neglected Tropical Diseases [38].

## Material and methods

### Ethical statement

Ethical approval was obtained from the Comité Bioética de la Facultad de Medicina (approved on 30/10/2020), Universidad Mayor de San Simón, Bolivia and from the Research Ethics Committee (REC) Instituto de Salud de Carlos III, Spain (CEI PI 74_2020)

### Overall Workflow

We introduce a comprehensive end-to-end pipeline that spans from sample digitization to model deployment. As illustrated in **Figure 1**, once the samples were collected and prepared, they were digitized using a smartphone application. The digitized images were subsequently uploaded to a telemedicine platform for labeling, creating the necessary input data for AI model training. Following this, multiple AI algorithms were trained and evaluated. Ultimately, the most effective AI models are deployed on edge devices to assist in the diagnostic process.

**Figure 1:**
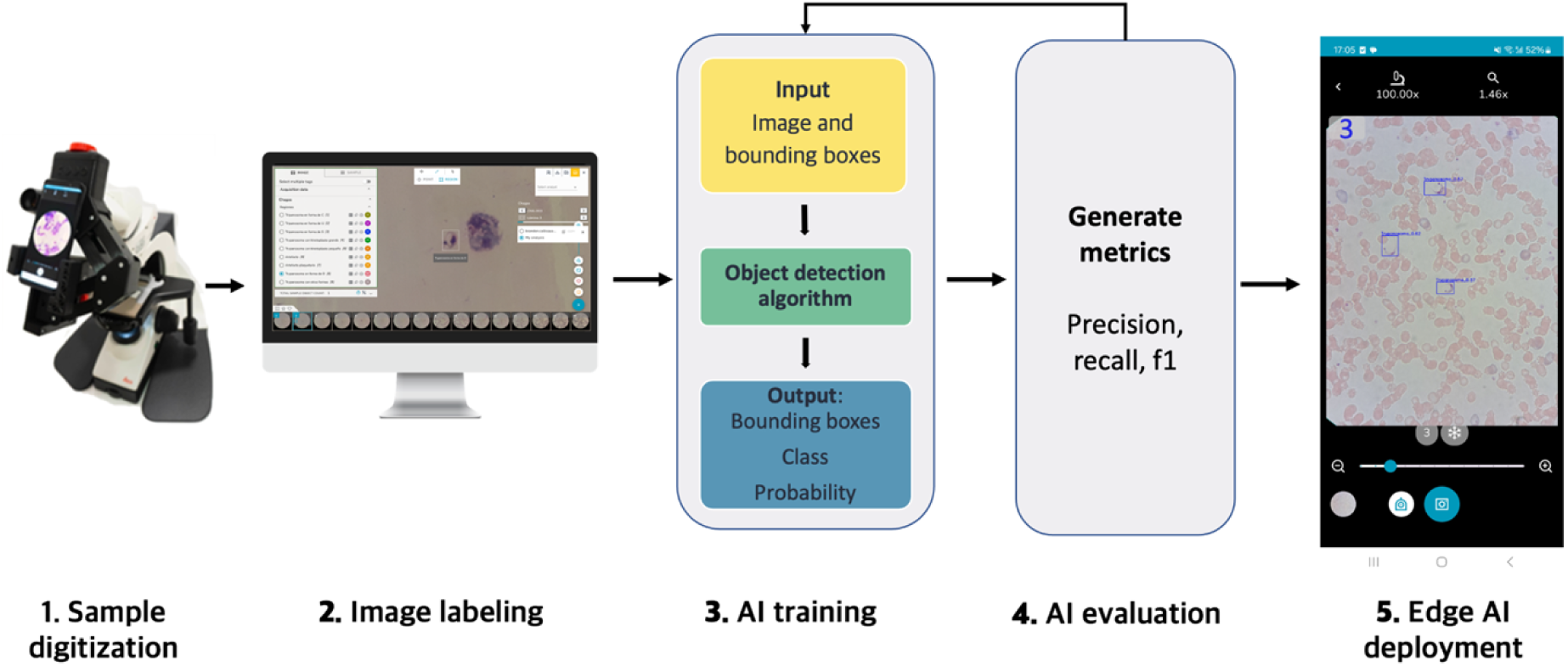
Simplified study design and workflow.

### Dataset

Our dataset comprises two subsets: we created a new dataset (Laboratorios de Investigación Médica (LABIMED) dataset) consisting of human thick and thin blood smears and cerebrospinal fluid smears (CSF), and a publicly available dataset containing mice thin blood smears (**Figure 2**). Integrating these diverse samples enhances the model’s exposure to clinical and experimental variability, simulating real-world diagnostic challenges and improving its adaptability across scenarios, as the sample preparation, staining protocol and imaging condition may differ from each laboratory. For instance, human thick blood smears retain multiple blood cell layers, whereas thin smears provide single-cell layers, altering parasite morphology and background noise. Similarly, staining methods affect color contrast and parasite visibility. Variations in imaging equipment - such as microscope resolution, lighting conditions, or camera - further compound these discrepancies.

**Figure 2:**
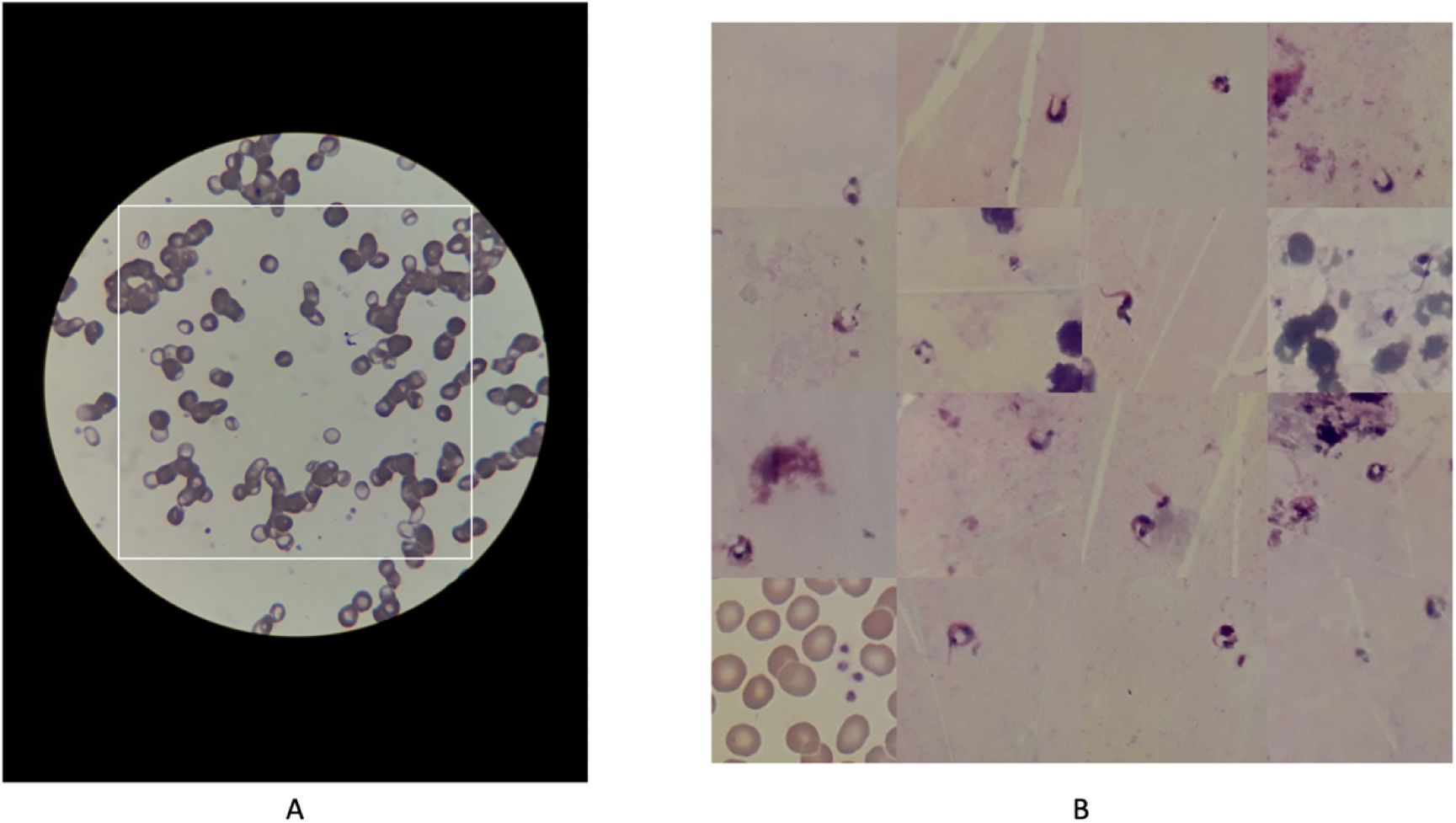
A) Field of view image, digitized with smartphone. B) Mosaic augmentation example.

A total of 36 smear preparations were collected from LABIMED, a reference laboratory for parasitology in Cochabamba, Bolivia, involving 9 positive subjects. Among these, 8 provided blood smear samples and 1 provided a cerebrospinal fluid (CSF) sample. All samples were stained with Giemsa. Specifically, 9 preparations from a single subject were CSF samples, 13 preparations from 4 subjects were thin blood smears, and 14 preparations from another 4 subjects were thick blood smears. The samples were digitized with Samsung Galaxy J7 prime, with 12-megapixel resolution together with the optical microscope, creating images of 4128×3096 pixels. Digitization was made using a 3D printed adapter coupled into the microscope, allowing the use of conventional microscopes to collect the images. Both positive and negative images were captured, resulting in 726 images.

The public dataset comprises 33 mice thin blood smear samples, totaling 675 images [31]. All images were Giemsa-stained and digitized using a smartphone camera coupled to an optical microscope with a 100x objective. Most images were captured at a resolution of 3,456 × 4,608 pixels, though a subset (2,448 × 3,264 pixels) reflects adjustments in camera settings during acquisition.

### Image labeling

For the development and evaluation of our object detection algorithm for *Trypanosoma cruzi*, accurate image labeling was crucial. We started by labeling the LABIMED dataset from scratch. Bounding boxes were placed around the identified parasite, indicating the location and the class of the parasite. In addition, some artifacts that look similar to the parasite were annotated. A total of 931 parasites were identified and annotated. Then, we trained a preliminary model to generate predictions on Morais *et al.* dataset, these predictions were then reviewed and refined by a human annotator, who adjusted the bounding boxes to ensure they accurately aligned with the parasites and corrected any false positives. This iterative process led to the creation of 3,151 labeled objects.

The labeled dataset was carefully split into two independent sets at sample preparation level for LABIMED dataset. This approach ensured that all images from the same preparation belong to the same dataset, whether used for training the AI model or validating its performance. The split was carried out after labeling the images to guarantee that each sample preparation type is presented in both the training and validation sets. The split was performed randomly, striving to achieve an 80%-20% split between the two sets. The result distribution is presented in **Table 1**.

**Table 1.** Dataset distribution of the training and validation algorithm.

| Sample type | Training |  |  | Validation |  |  | Total |  |  |
| --- | --- | --- | --- | --- | --- | --- | --- | --- | --- |
|  | #sample | #image | #label | #sample | #image | #label | #sample | #image | #label |
| Human CSF | 8 | 261 | 512 | 1 | 68 | 191 | 9 | 329 | 703 |
| Human Blood thick | 5 | 61 | 55 | 9 | 26 | 14 | 14 | 87 | 69 |
| Human Blood thin | 7 | 156 | 95 | 6 | 154 | 64 | 13 | 310 | 159 |
| Mice Blood thin | - | 570 | 2648 | - | 105 | 503 | - | 675 | 3151 |
| Total | 20 | 1048 | 3310 | 17 | 353 | 772 | 37 | 1401 | 4082 |

### Data preprocessing

Given the alignment of the smartphone with the microscope eyepiece, the area captured by the mobile phone was limited to a circular region, as illustrated in **Figure 3**. As the parasite size is very small, typically occupying an area of only 100×100 pixels within an image that is 4128×3096 pixels in size, to exclude non-informative regions, such as the black areas outside the circular field of view, we opted to use square images. To increase the relative size of the parasites within the image, we selected square images that encompass the entire field of view, as represented by the white square in **Figure 2**. This approach ensures that the visualized area is maximized while focusing on the relevant regions for analysis.

**Figure 3:**
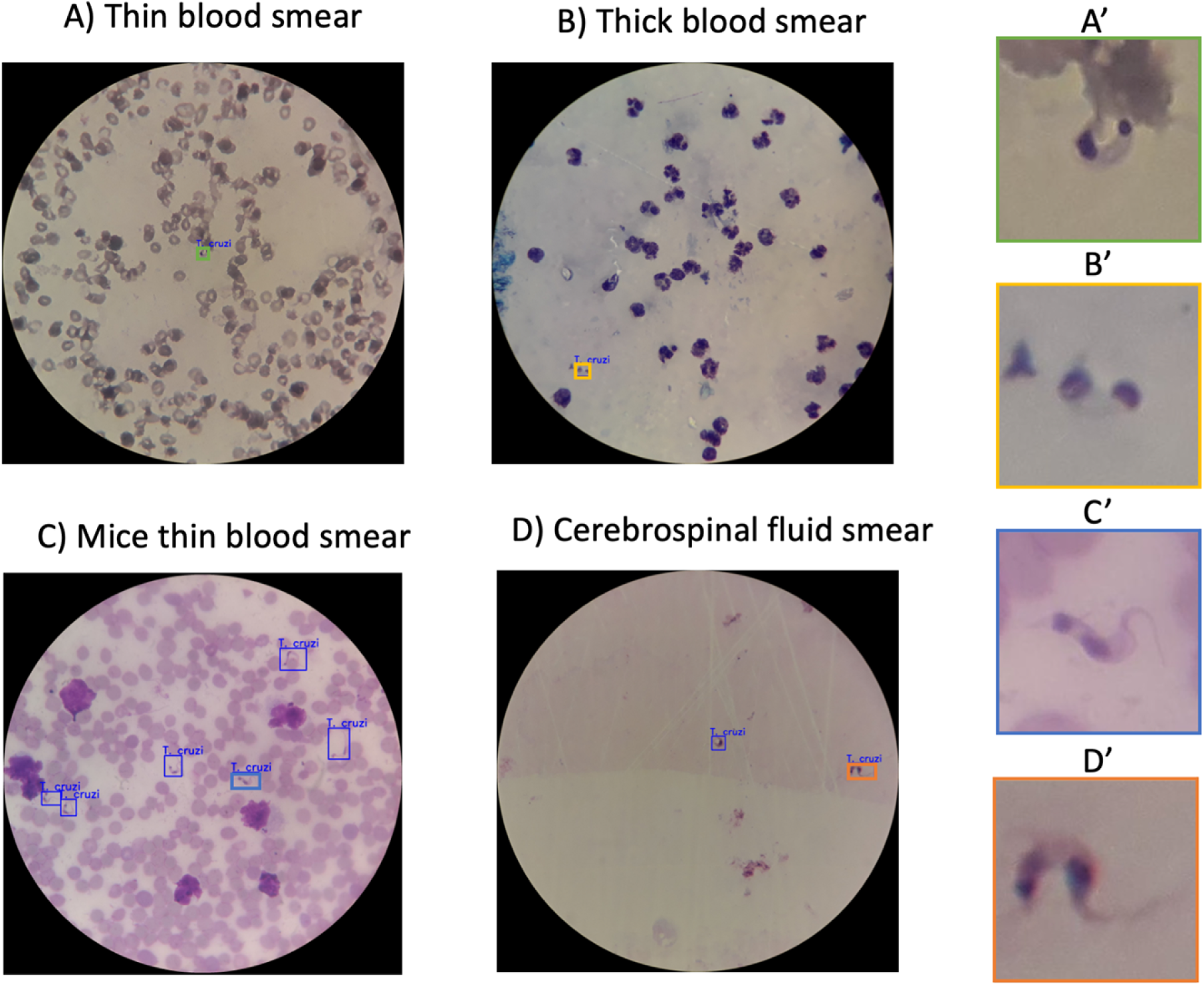
Microscopic images of blood and cerebrospinal fluid samples highlighting field of view and parasite visualization. The left panels display the full field of view for each sample type, while the right panels provide close-up views of parasites identified within the samples. Specifically, **A’** shows a parasite from the thin blood smear (Panel A), **B’** depicts a parasite from the thick blood smear (Panel B), **C’** presents a parasite from the mice thin blood smear (Panel C), and **D’** highlights a parasite from the cerebrospinal fluid smear (Panel D).

In addition, to overcome the problem of imbalance foreground and background in object detection tasks, we manually increased the number of parasites by applying mosaic augmentation, which consisted of cropping a 320×320 pixel patches that contained at least one parasite, and blending 4 images to create a new image of 640 x 640 pixel. This augmentation was only applied for the training set.

### AI algorithm

Object detection models can be divided into two groups: single stage detectors and two stage detectors. On the one hand, single stage detection algorithms are designed to make predictions within a single pass over the image which makes them faster but less accurate since they do not use a region proposal stage. On the other hand, two stage detection algorithms are more complex since they have a module that uses a CNN to propose regions of interest that are later refined in a second stage that performs the classification and localization. This process is usually more accurate but due to their nature, they are slower and require more computational resources.

In our work we focus on optimizing single-stage detectors to balance the need for speed and accuracy-as we want the AI model to be run in a mid-range Android device in real time. This approach ensures that real-time detection capabilities are maintained. In particular, we evaluated two different algorithms that represent the two main approaches for single stage detectors: Single Shot Detection (SSD) and YOLO [39–41].

The SSD-MobileNetV2 model integrates the SSD framework with the lightweight MobileNetV2 architecture. SSD predicted bounding boxes and class probabilities directly from feature maps at different scales in a single pass, enabling efficient object detection. MobileNetV2, designed for mobile optimization, employs depthwise separable convolutions to minimize computational costs, making it ideal for deployment on resource-constrained devices like smartphones. This combination strikes a balance between speed and accuracy. We initialized the model using pretrained weights from the COCO image dataset. For optimizing the loss function, we employed the RMSprop optimizer with an exponential decay learning rate, starting with a base learning rate of 0.008.

Conversely, YOLOv8 is one of the latest advancements in the YOLO series. It utilizes CSPDarknet53 as its backbone for feature extraction and incorporates a path aggregation network to enhance information flow across scales. YOLOv8 also introduces an anchor-free detection mechanism that predicts object centers directly, reducing the need for numerous box predictions and thus accelerating detection. The YOLOv8 family includes various variants, such as Nano, Small, Large, and Extra Large, each balancing accuracy and inference speed. For our application, we selected YOLOv8-Small for its optimal balance of these factors. The models were trained using the AdamW optimizer, configured with a learning rate of 0.002, a first momentum parameter of 0.9, and a second momentum parameter of 0.999. We also initialized the model using pretrained weights from the COCO image dataset for comparison purposes.

For both models, input size of 640×640 was used. During training, both models were subjected to on-the-fly data augmentation, including random flips, rotations, crops, and color jittering, to enhance generalization. Additionally, early stopping was employed to prevent overfitting, ensuring robust model performance.

For comparison purposes, we also trained a two-stage detector, Faster RCNN even though it is not suitable for smartphone deployment due to its higher computational demands.

### AI algorithm deployment

The trained AI model was exported to tflite format, with a size of approximately 11 megabytes for SSD Mobilenet and 45MB for YOLOv8 model. These models can be executed on Smartphones in real time. In addition, we created a customized Android application to execute the model.

## Results

Following data preprocessing, our training set comprised 7,249 augmented images with 66,557 annotated parasites, while the validation set included 1,099 images containing 1,668 labeled targets to assess model generalizability. After training, the top-performing model checkpoint (selected based on validation performance) was used to analyze validation images, successfully identifying *T. cruzi* parasites in diverse microscopy samples as visualized in **Figure 3**.

To evaluate the performance of our models, we used precision, recall, and F1 score as metrics. For each detection proposed by the algorithm, if the probability score of the detected object exceeds the threshold value and the Intersection over Union (IoU) with the ground truth bounding box is greater than 0.3, the detection is classified as a true positive. If the detected box does not overlap with any ground truth bounding box, it is classified as a false positive. Conversely, if an object is not detected by the algorithm, it is classified as a false negative. Precision, recall, and F1 score are then defined as follows:

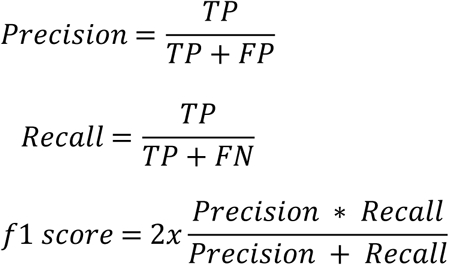

The performance of the models is summarized in **Table 2**. SSD-MobileNetV2 demonstrated superior performance on human samples, achieving a precision of 86%, a recall of 87%, and an F1 score of 86.5%. In comparison, YOLOv8-Small recorded precision, recall, and F1 scores of 83.6%, 87%, and 80.5%, respectively. Conversely, YOLOv8-Small outperformed SSD-MobileNetV2 on mice samples, attaining precision, recall, and F1 scores of 93.7%, 91.3%, and 92.5%, while SSD-MobileNetV2 achieved 95.8%, 85%, and 90.1%, respectively. Notably, the two-stage detector Faster R-CNN exhibited higher performance than the single-stage detectors, with precision, recall, and F1 scores of 93.6%, 81.4%, and 87.1% on human samples, and 96.8%, 90.3%, and 93.5% on mice samples.

**Table 2.** Model performance and inference time on Smartphone (Oppo Reno 6) with intel i5 of the three AI models for both human and mice samples.

| Architecture | Human |  |  | Mice |  |  | Inference on smartphone (milliseconds) |
| --- | --- | --- | --- | --- | --- | --- | --- |
|  | Precision | Recall | F1 score | Precision | Recall | F1 score |  |
| SSD-MobileNet v2 | 86 | 87 | 86.5 | 95.8 | 85 | 90.1 | 350 |
| YOLOv8-Small | 83.6 | 87 | 80.5 | 93.7 | 91.3 | 92.5 | 350 |
| Faster R-CNN (ResNet 50) | 93.6 | 81.4 | **87.1** | 96.8 | 90.3 | **93.5** | - |

When analyzing performance across different sample types, the models showed optimal results for mice with thin blood smears, achieving a precision of 95.8%, a recall of 85%, and an F1 score of 90.1%. In contrast, the performance on human thick blood smears was lower, with precision, recall, and F1 scores of 69.2%, 64.3%, and 66.7%, respectively. Performance for human thick smears drops compared to human CSF, thin, or mouse smears due to fewer training images and labels. These detailed results are presented in Table 3.

**Table 3:**
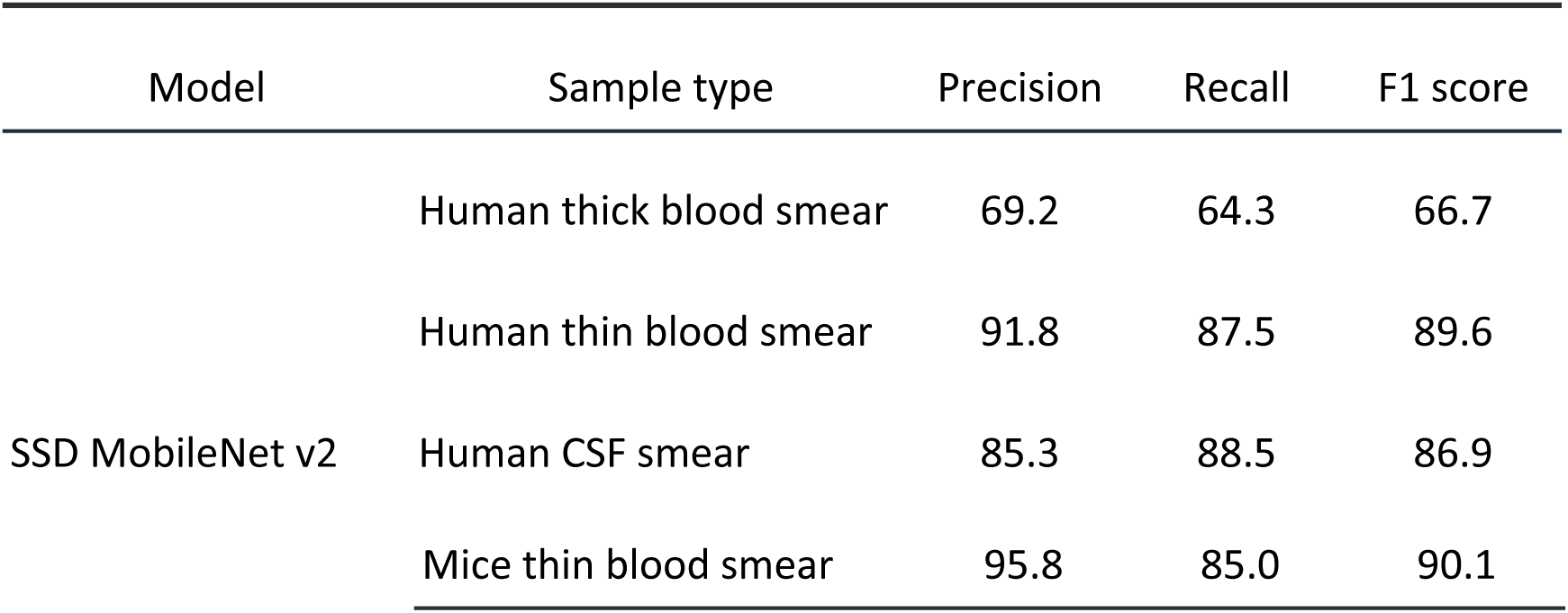
Performance of SSD MobileNet v2 separated by sample type.

## Discussion and conclusion

In this work we presented an end-to-end pipeline from image digitalization to image AI algorithm creation and smartphone deployment to automatically detect *T. cruzi* parasites. Laboratory diagnosis plays a crucial role in the control and monitoring of diseases. In the case of Chagas disease, the *T. cruzi* parasites detection in microscopic images remains a widely used method for acute phase diagnosis. Within endemic areas, in specialized and clinical laboratories, during the manual leukocyte count in blood smears or *Plasmodium* thick blood smear examination, *T. cruzi* trypomastigotes could be visualized. In experimental settings, such as drug discovery, automated detection of *T. cruzi* in microscopy images is essential for efficiently assessing treatment efficacy. Mice models are commonly used to evaluate potential drugs by monitoring parasitemia levels before and after treatment. Manual counting of parasites is time-consuming and prone to human error, which can introduce variability in results. Automated detection streamlines this process, providing rapid, consistent, and objective measurements of parasite burden. This enables researchers to accurately track disease progression and therapeutic response, ultimately facilitating the development of more effective treatments for Chagas disease [42–44]. Although various studies have explored automating parasite detection, achieving promising results but still facing certain limitations. Many algorithms operate on small image patches, cropping field-of-view images into numerous small areas, which increases computational time [31,32,35,36]. Alternatively, segmentation algorithms such as U-Net have been employed, offering detailed pixel-level information about the shape and size of parasites [33,37]. While segmentation provides precise measurements, it requires extensive pixel-level labeling, which is more time-consuming than bounding box annotations. Moreover, in the experimental contexts, the primary goal is to count the number of parasites rather than to determine their exact shape. Therefore, object detection, which focuses on identifying and counting objects within an image, could be more suitable for these purposes. In the majority of previous studies, the use of mice samples without including human samples, introducing possible bias.

In response to the limitations observed in existing methods, our study introduces an object detection approach specifically designed for Chagas disease diagnostics. We utilize advanced single-stage detectors, namely SSD-MobileNetV2 and YOLOv8, to enhance the detection process for deployment on smartphones. Our approach benefits from a comprehensive dataset that includes various sample types and incorporates robust data augmentation techniques, ensuring effective generalization across diverse imaging conditions. Our approach leverages a diverse dataset and robust data augmentation to ensure effective generalization across imaging conditions. Both SSD-MobileNetV2 and YOLOv8-Small demonstrated strong performance, achieving metrics consistently above 80% for all sample types, with SSD-MobileNetV2 excelling in human samples. The SSD-MobileNetV2 model’s inference time is approximately 350 milliseconds on a smartphone, underscoring its efficiency and suitability for real-time mobile diagnostics. By enabling real-time inference, our approach significantly enhances the practicality of AI-assisted Chagas disease diagnosis in field settings, where internet connectivity and computational resources are limited. Immediate and automated parasite detection reduces reliance on trained experts, minimizes diagnostic delays, and improves accessibility to timely diagnosis.

Even though Faster R-CNN is not suitable for deployment on smartphones due to its computational demands, the model can still be effectively utilized in a cloud-based environment. This allows for its integration into a human-in-the-loop labeling process, where it can assist in automating the initial labeling of data. By leveraging the model’s high accuracy, this approach can significantly reduce the time and effort required for manual labeling, while still allowing human oversight to ensure the quality of the annotations.

The primary limitation of our study is the sample size, particularly for human thick blood smears, where only 87 images with 69 labeled instances were available. This limited dataset likely contributed to the suboptimal performance for this sample type. We think this is due to the differences in parasite appearance and background characteristics between thick and thin smears, which affect the model’s ability to generalize across sample types. We believe that increasing the number of training samples could significantly enhance model performance, as evidenced by the strong results achieved with human thin blood smears, human CSF smears, and mice thin blood smears, where precision, recall, and F1 scores all exceeded 80%. Future work should focus on expanding the dataset, particularly for thick blood smears.

Additionally, microscopy has the potential to identify a wide range of parasites and cellular structures within the same image. Therefore, the AI should not be limited to detecting *T. cruzi* alone but should aim to function as a universal detector capable of identifying all discernible structures, such as malaria parasites, leishmania amastigotes, and cells, akin to a human microscopist. Furthermore, as multi-modal AI technology advances, these types of platforms have the potential to incorporate additional data sources, such as clinical and genetic data, a new generation of AI-driven diagnostics [45].

In conclusion, the proposed system offers real-time assistance in the diagnosis of Chagas disease by transforming a conventional light microscope into an intelligent point-of-care device. Our integrated telemedicine platform not only facilitates image storage and remote consultations but also streamlines the process of rapid image labeling and AI model development. Overall, new advancements in AI in medicine have the potential to provide accessible diagnostics for a wide range of infectious diseases, enhancing diagnostic accuracy and supporting global efforts to achieve the WHO’s targets for combating NTDs [38].

## Data Availability Statement

The microscopy image dataset, along with its associated annotations and the model, will be made publicly available.

## Acknowledgements

This project has received funding from the European Union’s Horizon 2020 research and innovation programme under grant agreement No 881062. LL was supported by a predoctoral grant IND2019/TIC-17167 (Comunidad de Madrid). This work was partially supported by the Bill and Melinda Gates Foundation (grant number Edge-Spot project INV-051355).

## Author contributions

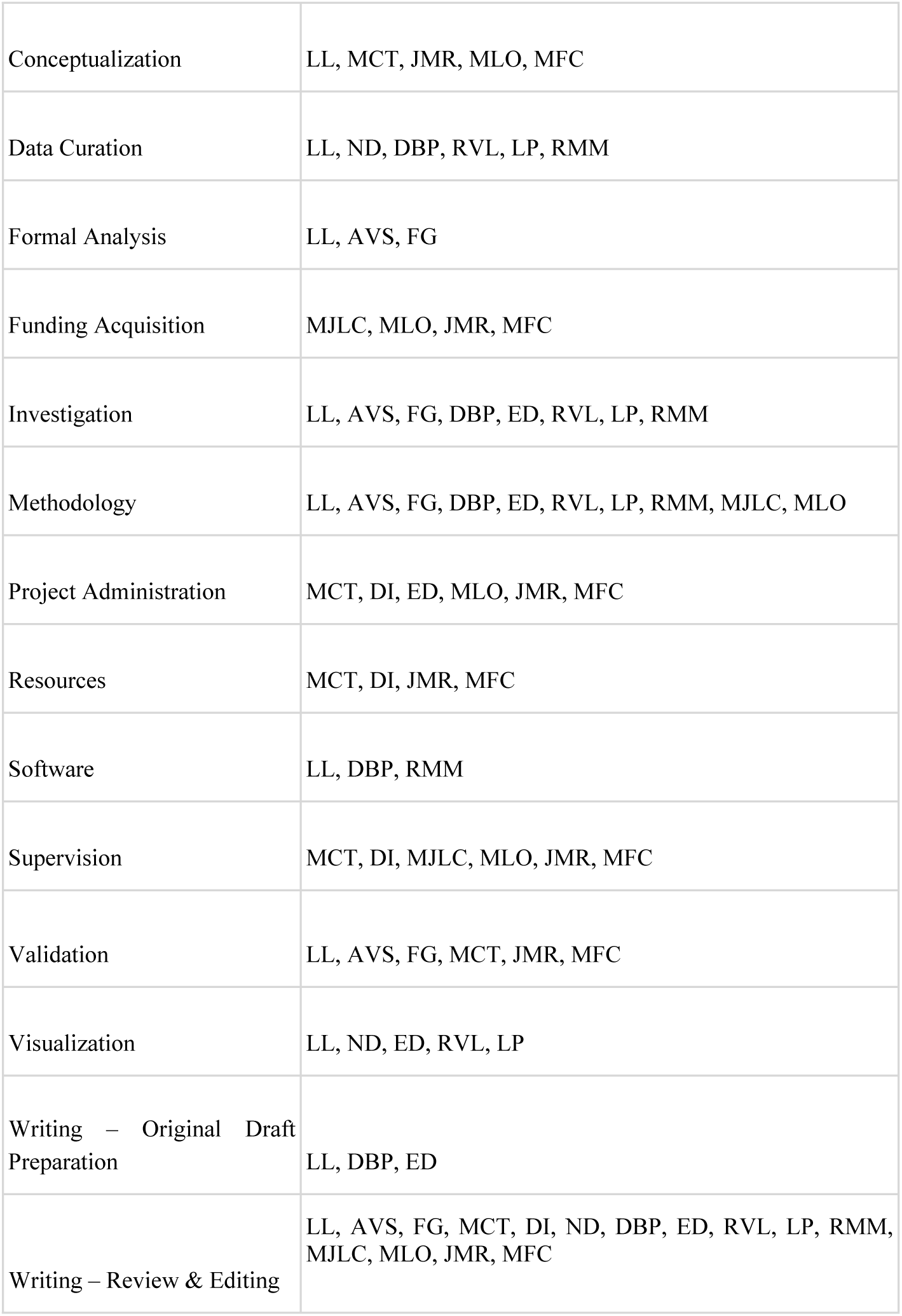

## Bibliography

1. World Health Organization. Chagas disease. [cited 22 Jul 2024]. Available: https://www.who.int/news-room/fact-sheets/detail/chagas-disease-(american-trypanosomiasis)

2. World Health Organization and others. Working to overcome the global impact of neglected tropical diseases: first WHO report on neglected tropical diseases. Savioli L, Daumerie D, editors. 2010.

3. Clark EH, Messenger LA, Whitman JD, Bern C. Chagas disease in immunocompromised patients. Clin Microbiol Rev. 2024;37: e0009923. doi:10.1128/cmr.00099-23

4. Clark EH, Bern C. Chagas Disease in People with HIV: A Narrative Review. Trop Med Infect Dis. 2021;6. doi:10.3390/tropicalmed6040198

5. Bhattacharyya T, Murphy N, Miles MA. Diversity of Chagas disease diagnostic antigens: Successes and limitations. PLoS Negl Trop Dis. 2024;18: e0012512. doi:10.1371/journal.pntd.0012512

6. Perez F, Vermeij D, Salvatella R, Castellanos LG, de Sousa AS. The use of rapid diagnostic tests for chronic Chagas disease: An expert meeting report. PLoS Negl Trop Dis. 2024;18: e0012340. doi:10.1371/journal.pntd.0012340

7. Schijman AG, Alonso-Padilla J, Britto C, Herrera Bernal CP. Retrospect, advances and challenges in Chagas disease diagnosis: a comprehensive review. Lancet Reg Health Am. 2024;36: 100821. doi:10.1016/j.lana.2024.100821

8. Suárez C, Nolder D, García-Mingo A, Moore DAJ, Chiodini PL. Diagnosis and Clinical Management of Chagas Disease: An Increasing Challenge in Non-Endemic Areas. Res Rep Trop Med. 2022;13: 25–40. doi:10.2147/RRTM.S278135

9. Lopez-Albizu C, Rivero R, Ballering G, Freilij H, Santini MS, Bisio MMC. Laboratory diagnosis of Trypanosoma cruzi infection: a narrative review. Front Parasitol. 2023;2. doi:10.3389/fpara.2023.1138375

10. Teixeira de Sousa DR, de Oliveira Guerra JA, Ortiz JV, do Nascimento Couceiro K, da Silva E Silva MRH, Jorge Brandão AR, et al. Acute Chagas disease associated with ingestion of contaminated food in Brazilian western Amazon. Trop Med Int Health. 2023;28: 541–550. doi:10.1111/tmi.13899

11. Monteiro WM, Barbosa M das GV, Guerra JA de O, Melo GC de, Barbosa LRA, Machado KVA, et al. Driving forces for strengthening the surveillance of Chagas disease in the Brazilian Amazon by “training the eyes” of malaria microscopists. Rev Soc Bras Med Trop. 2020;53: e20190423. doi:10.1590/0037-8682-0423-2019

12. Muehlematter UJ, Daniore P, Vokinger KN. Approval of artificial intelligence and machine learning-based medical devices in the USA and Europe (2015-20): a comparative analysis. Lancet Digit Health. 2021;3: e195–e203. doi:10.1016/S2589-7500(20)30292-2

13. Benjamens S, Dhunnoo P, Meskó B. The state of artificial intelligence-based FDA-approved medical devices and algorithms: an online database. npj Digital Med. 2020;3:118. doi:10.1038/s41746-020-00324-0

14. Xu H, Usuyama N, Bagga J, Zhang S, Rao R, Naumann T, et al. A whole-slide foundation model for digital pathology from real-world data. Nature. 2024;630: 181–188. doi:10.1038/s41586-024-07441-w

15. Vorontsov E, Bozkurt A, Casson A, Shaikovski G, Zelechowski M, Severson K, et al. A foundation model for clinical-grade computational pathology and rare cancers detection. Nat Med. 2024;30: 2924–2935. doi:10.1038/s41591-024-03141-0

16. Fan BE, Yong BSJ, Li R, Wang SSY, Aw MYN, Chia MF, et al. From microscope to micropixels: A rapid review of artificial intelligence for the peripheral blood film. Blood Rev. 2023; 101144. doi:10.1016/j.blre.2023.101144

17. Wang W, Luo M, Guo P, Wei Y, Tan Y, Shi H. Artificial intelligence-assisted diagnosis of hematologic diseases based on bone marrow smears using deep neural networks. Comput Methods Programs Biomed. 2023;231: 107343. doi:10.1016/j.cmpb.2023.107343

18. Feng R, Li S, Zhang Y. AI-powered microscopy image analysis for parasitology: integrating human expertise. Trends Parasitol. 2024;40: 633–646. doi:10.1016/j.pt.2024.05.005

19. Elmehankar M, Etewa S, Metwally A. Artificial intelligence applications in medical parasitology: A comprehensive review. Egyptian Academic Journal of Biological Sciences, E Medical Entomology & Parasitology. 2023;15: 35–43. doi:10.21608/eajbse.2023.322464

20. Das D, Vongpromek R, Assawariyathipat T, Srinamon K, Kennon K, Stepniewska K, et al. Field evaluation of the diagnostic performance of EasyScan GO: a digital malaria microscopy device based on machine-learning. Malar J. 2022;21: 122. doi:10.1186/s12936-022-04146-1

21. Mujahid M, Rustam F, Shafique R, Montero EC, Alvarado ES, de la Torre Diez I, et al. Efficient deep learning-based approach for malaria detection using red blood cell smears. Sci Rep. 2024;14: 13249. doi:10.1038/s41598-024-63831-0

22. Horning MP, Delahunt CB, Bachman CM, Luchavez J, Luna C, Hu L, et al. Performance of a fully-automated system on a WHO malaria microscopy evaluation slide set. Malar J. 2021;20: 110. doi:10.1186/s12936-021-03631-3

23. Oyibo P, Jujjavarapu S, Meulah B, Agbana T, Braakman I, van Diepen A, et al. Schistoscope: An Automated Microscope with Artificial Intelligence for Detection of Schistosoma haematobium Eggs in Resource-Limited Settings. Micromachines (Basel). 2022;13. doi:10.3390/mi13050643

24. Holmström O, Linder N, Ngasala B, Mårtensson A, Linder E, Lundin M, et al. Point-of-care mobile digital microscopy and deep learning for the detection of soil-transmitted helminths and Schistosoma haematobium. Glob Health Action. 2017;10:1337325. doi:10.1080/16549716.2017.1337325

25. Armstrong M, Harris AR, D’Ambrosio MV, Coulibaly JT, Essien-Baidoo S, Ephraim RKD, et al. Point-of-Care Sample Preparation and Automated Quantitative Detection of Schistosoma haematobium Using Mobile Phone Microscopy. Am J Trop Med Hyg. 2022;106: 1442–1449. doi:10.4269/ajtmh.21-1071

26. Meulah B, Bengtson M, Lieshout LV, Hokke CH, Kreidenweiss A, Diehl J-C, et al. A review on innovative optical devices for the diagnosis of human soil-transmitted helminthiasis and schistosomiasis: from research and development to commercialization. Parasitology. 2023;150: 137–149. doi:10.1017/S0031182022001664

27. Lundin J, Suutala A, Holmström O, Henriksson S, Valkamo S, Kaingu H, et al. Diagnosis of soil-transmitted helminth infections with digital mobile microscopy and artificial intelligence in a resource-limited setting. PLoS Negl Trop Dis. 2024;18: e0012041. doi:10.1371/journal.pntd.0012041

28. Ward P, Dahlberg P, Lagatie O, Larsson J, Tynong A, Vlaminck J, et al. Affordable artificial intelligence-based digital pathology for neglected tropical diseases: A proof-of-concept for the detection of soil-transmitted helminths and Schistosoma mansoni eggs in Kato-Katz stool thick smears. PLoS Negl Trop Dis. 2022;16: e0010500. doi:10.1371/journal.pntd.0010500

29. Dacal E, Bermejo-Peláez D, Lin L, Álamo E, Cuadrado D, Martínez Á, et al. Mobile microscopy and telemedicine platform assisted by deep learning for the quantification of Trichuris trichiura infection. PLoS Negl Trop Dis. 2021;15: e0009677. doi:10.1371/journal.pntd.0009677

30. Lin L, Dacal E, Díez N, Carmona C, Martin Ramirez A, Barón Argos L, et al. Edge Artificial Intelligence (AI) for real-time automatic quantification of filariasis in mobile microscopy. PLoS Negl Trop Dis. 2024;18: e0012117. doi:10.1371/journal.pntd.0012117

31. Morais MCC, Silva D, Milagre MM, de Oliveira MT, Pereira T, Silva JS, et al. Automatic detection of the parasite Trypanosoma cruzi in blood smears using a machine learning approach applied to mobile phone images. PeerJ. 2022;10: e13470. doi:10.7717/peerj.13470

32. Pereira A, Pyrrho A, Vanzan D, Mazza L, Gomes JG. Deep Convolutional Neural Network applied to Chagas Disease Parasitemia Assessment. Anais do 14 Congresso Brasileiro de Inteligência Computacional. ABRICOM; 2020. pp. 1–8. doi:10.21528/CBIC2019-119

33. Sanchez-Patino N, Toriz-Vazquez A, Hevia-Montiel N, Perez-Gonzalez J. Convolutional neural networks for chagas’ parasite detection in histopathological images. Annu Int Conf IEEE Eng Med Biol Soc. 2021;2021: 2732–2735. doi:10.1109/EMBC46164.2021.9629563

34. Soberanis-Mukul R, Uc-Cetina V, Brito-Loeza C, Ruiz-Piña H. An automatic algorithm for the detection of Trypanosoma cruzi parasites in blood sample images. Comput Methods Programs Biomed. 2013;112: 633–639. doi:10.1016/j.cmpb.2013.07.013

35. Uc-Cetina V, Brito-Loeza C, Ruiz-Piña H. Chagas Parasite Detection in Blood Images Using AdaBoost. Comput Math Methods Med. 2015;2015: 139681. doi:10.1155/2015/139681

36. Jung T, Anzaku ET, Özbulak U, Magez S, Van Messem A, De Neve W. Automatic Detection of Trypanosomosis in Thick Blood Smears Using Image Pre-processing and Deep Learning. In: Singh M, Kang D-K, Lee J-H, Tiwary US, Singh D, Chung W-Y, editors. Intelligent human computer interaction: 12th international conference, IHCI 2020, daegu, south korea, november 24–26, 2020, proceedings, part II. Cham: Springer International Publishing; 2021. pp. 254–266. doi:10.1007/978-3-030-68452-5_27

37. Ojeda-Pat A, Martin-Gonzalez A, Soberanis-Mukul R. Convolutional Neural Network U-Net for Trypanosoma cruzi Segmentation. In: Brito-Loeza C, Espinosa-Romero A, Martin-Gonzalez A, Safi A, editors. Intelligent computing systems: third international symposium, ISICS 2020, sharjah, united arab emirates, march 18–19, 2020, proceedings. Cham: Springer International Publishing; 2020. pp. 118–131. doi:10.1007/978-3-030-43364-2_11

38. World Health Organization, editor. Ending the neglect to attain the Sustainable Development Goals: a road map for neglected tropical diseases 2021–2030. World Health Organization; 2021.

39. Liu W, Anguelov D, Erhan D, Szegedy C, Reed S, Fu C-Y, et al. SSD: Single shot multibox detector. In: Leibe B, Matas J, Sebe N, Welling M, editors. European Conference on Computer VIsion (ECCV). Cham: Springer International Publishing; 2016. pp. 21–37. doi:10.1007/978-3-319-46448-0_2

40. Redmon J, Divvala S, Girshick R, Farhadi A. YOLO You Only Look Once: Unified, Real-Time Object Detection. 2016 IEEE Conference on Computer Vision and Pattern Recognition (CVPR). IEEE; 2016. pp. 779–788. doi:10.1109/CVPR.2016.91

41. Jocher G, Qiu J, Chaurasia A. Ultralytics YOLO. 2023.

42. Álvarez MG, Hernández Y, Bertocchi G, Fernández M, Lococo B, Ramírez JC, et al. New Scheme of Intermittent Benznidazole Administration in Patients Chronically Infected with Trypanosoma cruzi: a Pilot Short-Term Follow-Up Study with Adult Patients. Antimicrob Agents Chemother. 2016;60: 833–837. doi:10.1128/AAC.00745-15

43. Gabaldón-Figueira JC, Martinez-Peinado N, Escabia E, Ros-Lucas A, Chatelain E, Scandale I, et al. State-of-the-Art in the Drug Discovery Pathway for Chagas Disease: A Framework for Drug Development and Target Validation. Res Rep Trop Med. 2023;14: 1–19. doi:10.2147/RRTM.S415273

44. De Rycker M, Wyllie S, Horn D, Read KD, Gilbert IH. Anti-trypanosomatid drug discovery: progress and challenges. Nat Rev Microbiol. 2023;21: 35–50. doi:10.1038/s41579-022-00777-y

45. Rajpurkar P, Lungren MP. The current and future state of AI interpretation of medical images. N Engl J Med. 2023;388: 1981–1990. doi:10.1056/NEJMra2301725

